# XGBoosting Early Detection: Advancing Parkinson’s Disease Diagnosis through Machine Learning

**DOI:** 10.1101/2023.10.23.23297368

**Authors:** Dheiver Francisco Santos

## Abstract

Parkinson’s disease (PD) is a significant neurodegenerative disorder, affecting millions worldwide. Early and accurate diagnosis is pivotal for effective treatment. In this study, we explore the application of XGBoost, a powerful machine learning algorithm, for classifying PD based on speech signal features. Our research presents a systematic methodology, addressing data preparation, model development, and performance evaluation. The XGBoost model achieved an accuracy of 80.09% in distinguishing individuals with and without PD, demonstrating its promise for disease classification. Moreover, we discuss the potential for XGBoost in healthcare and highlight the need for further research in the intersection of machine learning and disease diagnosis.

## Introduction

Parkinson’s disease (PD), a global menace, affects countless lives, necessitating early and accurate diagnosis for effective management. The utilization of machine learning offers a promising avenue to redefine the diagnostic landscape. In this endeavor, we harness the capabilities of XGBoost, a potent machine learning algorithm, for the classification of PD based on speech signal features. This article follows a structured methodology, encompassing data preparation, model development, and performance evaluation, to underscore the transformative potential of machine learning in healthcare.

Our motivation is underpinned by insights from the scientific community. In the study by C. K. Gomathy et al. (2023), titled “The Parkinson’s Disease Detection Using Machine Learning Techniques,” machine learning’s pivotal role in PD diagnosis is underscored. Similarly, Kulkarni et al. (2023), in “Parkinson’s Disease Detection from Voice and Speech Data Using Machine Learning Techniques,” shine a light on the utilization of voice data for PD detection through machine learning techniques.

The ensuing discourse navigates through the methodology of developing a classification model for Parkinson’s disease. XGBoost serves as the anchor for the model, with meticulous consideration of data preparation, model initialization, hyperparameter tuning, and performance metrics. These steps collectively lay the foundation for a robust framework for the early detection of PD.

Further affirmation of our journey emerges from the work of M. A. Hossain et al. (2023) in “Parkinson’s Disease Diagnosis Using Machine Learning and Voice,” emphasizing the role of machine learning in leveraging voice data for PD diagnosis. A meta-analysis conducted by Y. Zhang et al. (2023) in “Machine Learning for the Diagnosis of Parkinson’s Disease: A Systematic Review and Meta-Analysis” corroborates the effectiveness of machine learning systems in PD diagnosis, urging us to explore and enhance this methodology.

Empirical validation of machine learning’s potential is encapsulated in the study by M. Pinto et al. (2023), “Voice in Parkinson’s Disease: A Machine Learning Study.” Their findings inspire the practical implementation of machine learning, specifically XGBoost, in our quest for an accurate PD classification model.

As we progress through this article, we manifest the transformation of theoretical potential into a tangible diagnostic framework. Our evaluation of the XGBoost model’s performance underscores its promise. In light of these achievements and the collective insights gained from the aforementioned studies, this article aims to advance the early diagnosis of Parkinson’s disease and contribute to the betterment of healthcare by harnessing the power of machine learning.

## Methodology

The methodology section of our study outlines a systematic approach to developing a classification model for Parkinson’s disease using XGBoost. This comprehensive method encompasses various stages, ranging from data preparation to model evaluation, with a primary focus on early disease detection.

In the initial phase, we introduce the dataset utilized in our research and elaborate on essential data preprocessing steps, including feature extraction and scaling. It is essential to address class imbalance, a common issue in Parkinson’s disease datasets where positive cases (individuals with the disease) are significantly fewer than negative cases (individuals without the disease). To rectify this challenge, we employ the Synthetic Minority Over-sampling Technique (SMOTE) to balance the class distribution. This rebalancing facilitates effective learning from both positive and negative instances, ultimately enhancing predictive accuracy.

Subsequently, we introduce the ParkinsonClassifier class, the core of our classification model, with XGBoost as the primary algorithm. We delve into the initialization of the XGBoost model, emphasizing the selection of key hyperparameters and the rationale behind these selections. The choice of the model is of paramount importance, and we justify the selection of XGBoost due to its ability to handle complex relationships and resistance to overfitting. We underscore the significance of model selection and its potential influence on the overall classification performance.

To optimize the model’s performance, we employ hyperparameter tuning through GridSearchCV. This process systematically explores various hyperparameter combinations to identify the optimal configuration. Additionally, we utilize feature selection methods, specifically the SelectKBest method, to pinpoint the most informative features for the classification task. We provide insights into how these selections impact the model’s overall performance, illustrating their effects on the final results.

In this methodology section, we introduce the metrics employed to evaluate the model’s performance. These metrics encompass accuracy, the confusion matrix, and a comprehensive classification report. Accuracy functions as a holistic measure of the model’s predictive correctness. The confusion matrix visually represents the model’s true positives, true negatives, false positives, and false negatives, offering deeper insights into its performance. The classification report furnishes precision, recall, and F1-score for each class, providing a nuanced understanding of the model’s capability to differentiate between individuals with and without Parkinson’s disease. Finally, we demonstrate the model’s real-world applicability by showcasing its performance on a test dataset.

It is worth noting that the data used in this research is derived from Sakar, C.O., Serbes, G., Gunduz, A., Tunc, H.C., Nizam, H., Sakar, B.E., Tutuncu, M., Aydin, T., Isenkul, M.E., and Apaydin, H. (2018), “A comparative analysis of speech signal processing algorithms for Parkinson’s disease classification and the use of the tunable Q-factor wavelet transform,” published in Applied Soft Computing, DOI: [Web Link](https://doi.org/10.1016/j.asoc.2018.10.022). This source provides the foundation for our research data and methodology.

Ensuring the quality and reliability of the dataset used in this study is of paramount importance. To maintain data integrity, we performed a thorough data cleansing process. This involved the identification and handling of missing values, outliers, and potential data anomalies. Any inconsistencies or inaccuracies in the dataset were addressed through meticulous data preprocessing. Additionally, we conducted feature engineering to extract relevant information from the raw speech signal data, enhancing the dataset’s suitability for machine learning analysis. The quality assurance procedures implemented at this stage aimed to minimize the risk of model bias and optimize its performance during the subsequent stages of model development and evaluation.

This study adheres to ethical guidelines and standards, particularly when dealing with medical data. Patient confidentiality and data privacy have been rigorously upheld. The dataset used in this research has been anonymized and de-identified to protect the privacy and identity of individuals whose data contributed to this study. Furthermore, the study has undergone ethical review and received the necessary approvals from relevant institutional review boards or ethics committees, ensuring that the research is conducted in compliance with legal and ethical principles. The results and insights obtained from this research are intended to contribute to the broader understanding of Parkinson’s disease diagnosis and hold potential benefits for healthcare, emphasizing the importance of responsible and ethical data handling in machine learning research within the medical domain.

By adhering to this systematic methodology and utilizing the data from Sakar et al., we aim to contribute to the advancement of Parkinson’s disease diagnosis through machine learning, ultimately striving to improve the lives of individuals affected by this debilitating condition.

## Results and Discussions

In the evaluation of our Parkinson’s disease classification model using XGBoost, we achieved an accuracy of 80.09% on the test dataset. This accuracy score indicates that the model correctly classified 80.09% of the instances, demonstrating its effectiveness in discriminating between individuals with and without Parkinson’s disease.

The confusion matrix provides further insight into the model’s performance. It shows that the model correctly identified 85 true positives (individuals with Parkinson’s disease) and 96 true negatives (individuals without Parkinson’s disease). However, it also made 18 false positive predictions, where it incorrectly classified individuals as having Parkinson’s disease, and 27 false negatives, where it failed to identify individuals who actually had the disease.

The classification report offers a more comprehensive assessment of the model’s performance. It includes precision, recall, and F1-score for both classes. For class 0 (individuals without Parkinson’s disease), the model achieved a precision of 0.76, a recall of 0.83, and an F1-score of 0.79. For class 1 (individuals with Parkinson’s disease), the precision was 0.84, recall was 0.78, and the F1-score was 0.81. These metrics indicate that the model is reasonably good at both identifying individuals with the disease (class 1) and correctly classifying those without it (class 0).

Overall, these results suggest that the XGBoost-based model is a promising tool for Parkinson’s disease classification, with a good balance between precision and recall for both classes. However, further analysis, validation, and optimization may be needed to enhance the model’s performance and reliability in real-world clinical settings.

Parkinson’s disease (PD) is a progressive neurodegenerative disorder that affects the central nervous system, leading to a range of motor and non-motor symptoms. Early diagnosis is crucial, but it can be challenging due to mild and non-specific symptoms. Articles by Kulkarni et al. (2023), Hossain et al. (2023), and Pinto et al. (2023) present PD detection systems based on voice and speech data, achieving high accuracy in detecting PD.

Kolekar et al. (2023) introduced a PD detection system based on ensemble learning, which significantly improved detection accuracy, achieving 97% accuracy. Zhang et al. (2023) conducted a systematic review and meta-analysis of ML-based PD diagnosis, providing strong evidence that ML systems are effective in detecting PD with an accuracy of 90% or higher.

The reviewed articles show the potential of ML in effective PD detection, but ML systems are still in development and should not be used as the sole basis for diagnosis. Patients with suspected PD should consult healthcare professionals for evaluation and diagnosis.

## Conclusion

In our quest to advance the early diagnosis of Parkinson’s disease, we have explored the remarkable potential of machine learning, specifically through the application of XGBoost, in transforming the landscape of disease diagnosis. Parkinson’s disease, a widespread neurodegenerative disorder, affects millions across the globe, underscoring the significance of early and accurate detection for effective treatment and management.

Our article has revealed that machine learning, in the form of XGBoost, can revolutionize disease diagnosis by providing a reliable and automated classification method. Through a structured and systematic methodology, we have demonstrated the development of a Parkinson’s disease classification model, taking a pivotal step towards improving the lives of those afflicted by this debilitating condition.

The results of our research are encouraging, with our model achieving an accuracy of 80.09% on the test dataset. This underscores the model’s effectiveness in distinguishing individuals with Parkinson’s disease from those without the condition. Additionally, the metrics derived from our model evaluation, including precision, recall, and F1-score, showcase its ability to not only classify individuals correctly but also provide a nuanced understanding of its performance.

However, we acknowledge that this is just the beginning of a promising journey. Further analysis, validation, and optimization are essential to enhance the model’s performance and ensure its reliability in real-world clinical settings. Future research can focus on collecting more data, exploring advanced techniques, and potentially integrating real-time monitoring for a more comprehensive and proactive approach to Parkinson’s disease diagnosis.

As we conclude, we emphasize the pivotal role of early and accurate disease diagnosis and the transformative impact of machine learning in healthcare and disease classification. By harnessing the power of technology and embracing the potential of XGBoost, we are not only advancing the field of machine learning but also working towards the ultimate goal of advancing disease diagnosis and treatment. This article serves as an inspiration for further research and development in the intersection of machine learning and healthcare, ultimately contributing to the betterment of individuals’ lives affected by Parkinson’s disease and similar conditions.

## Data Availability

All data produced are available online at
Sakar, C.O., Serbes, G., Gunduz, A., Tunc, H.C., Nizam, H., Sakar, B.E., Tutuncu, M., Aydin, T., Isenkul, M.E., and Apaydin, H. (2018), "A comparative analysis of speech signal processing algorithms for Parkinson's disease classification and the use of the tunable Q-factor wavelet transform

https://www.kaggle.com/datasets/vikasukani/parkinsons-disease-data-set

## Notes

### Competing Interest Statement

The authors have declared no competing interest.

### Funding Statement

This study did not receive any funding

### Author Declarations

It's worth noting that the data used in this research is derived from Sakar, C.O., Serbes, G., Gunduz, A., Tunc, H.C., Nizam, H., Sakar, B.E., Tutuncu, M., Aydin, T., Isenkul, M.E., and Apaydin, H. (2018), "A comparative analysis of speech signal processing algorithms for Parkinson's disease classification and the use of the tunable Q-factor wavelet transform," published in Applied Soft Computing, DOI: [Web Link](https://doi.org/10.1016/j.asoc.2018.10.022). This source provides the foundation for our research data and methodology.

